# How well does spirometry capture the symptom range in primary ciliary dyskinesia? – a cross-sectional national study

**DOI:** 10.1101/2025.08.26.25334463

**Authors:** Yin Ting Lam, Mirjam Koller, Leonie D Schreck, Christian Clarenbach, Andreas Jung, Elisabeth Kieninger, Claudia E Kuehni, Myrofora Goutaki

**Author notes:** Corresponding Author: Myrofora Goutaki, Institute of Social and Preventive Medicine, University of Bern, Mittelstrasse 43, 3012 Bern, Switzerland.

## Abstract

**Introduction:** Patients with primary ciliary dyskinesia (PCD) present a variety of respiratory symptoms. Spirometry, particularly forced expiratory volume in 1 s (FEV1), is the most commonly used outcome measure in clinical follow-up, however, it is not known how well it captures the range of respiratory symptoms experienced by patients.

**Methods:** We sent the FOLLOW-PCD questionnaire to all patients ≥14 years and parents of children, registered in the Swiss PCD Registry, asking about upper and lower respiratory symptoms. In patients who had a routine lung function done within a year of survey completion, we extracted data from clinical records and calculated spirometric indices z-scores based on the Global Lung Function Initiative references. We used linear regression to study associations between FEV1, forced vital capacity (FVC), FEV1/FVC and frequency of respiratory symptoms, adjusted for age, sex, and regular physiotherapy.

**Results:** 64 out of 99 invited patients (67%) completed the survey; for 54 of them (median age 24 years, IQR 15-47; 50% females) we also had an FEV1 measurement (mean z-score - 2.29 [- 3.37 – −1.03]), with 2.2 months median time between survey and lung function test. Patients reporting wheeze (76%) had lower FEV1 and FEV1/FVC z-scores (FEV1 z-score for infrequent and frequent wheeze compared to no wheeze: −1.13 [95%CI −2.13 – −0.12] and −1.11 [−2.20 – - 0.20] respectively). Similarly patients reporting frequent shortness of breath (29.5%) had also lower FEV1 and FEV1/FVC z-scores (FEV1 z-score for frequent shortness of breath compared to no shortness of breath: −1.27 [−2.50 – −0.04]). We found no signs of association between reported nasal symptoms, snoring, cough, sputum production, and chest pain with FEV1, FVC or FEV1/FVC.

**Conclusion:** In our study, self-reported wheeze and frequent shortness of breath were associated with lower FEV1 and FEV1/FVC, commonly used for patient follow-up. However, we need additional outcome measures e.g., lung clearance index, imaging outcomes, or upper airway assessments, together with regular and standardised assessment of patient-reported symptoms, to capture the range of respiratory morbidity patients with PCD experience in daily life and guide management successfully.

## INTRODUCTION

Selecting the optimal assessment tools for clinical follow-up of chronic diseases is one of the main challenges healthcare professionals and researchers often face. Appropriate outcome measures are of outmost importance for research studies, especially clinical trials, but also in daily practice for the clinical follow-up of patients. Objective measures are often considered more reliable to capture disease progression and to allow comparisons over time. Over the past decades, however, patient-reported outcomes have gained importance as they offer a different perspective relating to the patients’ perception of disease severity and how it affects their daily lives [1]. This is especially relevant for rare or complex diseases with large phenotypic variability, such as primary ciliary dyskinesia (PCD) [2, 3].

PCD prevalence is estimated to be 1 in 7500 and symptoms start early in life and progress over the years [4]. Most patients develop recurrent upper and lower respiratory symptoms, leading to chronic rhinosinusitis, hearing impairment, and bronchiectasis, [5–7]. A scoping review showed that commonly used lung health outcome measures for PCD have not been validated, and reporting is not standardised [8]. For the upper airways and ears, the heterogeneity in outcome definitions or measures is even more prominent [9]. Several objective measures are available to capture different aspects of lung health in PCD, but spirometry remains the most used for all patients worldwide. It is recommended for regular follow-ups, usually every 3 to 6 months [10]. Spirometry is also commonly used in research to assess lung disease progression [11–14]. Measures such as multiple breath washout or structural and functional imaging modalities have shown better sensitivity in detecting early signs of disease and monitoring lung health; however, these are not commonly available or used in routine care [14–17]. Spirometry, specifically forced expiratory volume in 1s (FEV1) was the only objective measure together with microbiology, included in the recently published BEAT-PCD (Better Experimental Approaches to Treat Primary Ciliary Dyskinesia) consensus statement on a core outcome set for pulmonary disease interventions [18]. The core outcome set also included exacerbations and health-related quality of life as patient-reported outcomes, however these still are not used regularly or are designed specifically for use in research [19–21]. Since there is a lack of standardised and reliable clinical outcome measures that capture patient-reported symptoms, clinical decision making often relies mainly on objective measures, in particular spirometry. We aimed to study associations between respiratory symptoms and airway obstruction, assessed by spirometry among patients with PCD living in Switzerland. We hypothesised that spirometry alone would not be able to reflect the range of symptoms patients with PCD experience.

## METHODS

### Study design and study population

Our study is nested in the Swiss PCD registry (CH-PCD), a population-based patient registry (www.clinicaltrials.gov identifier <u>NCT03606200</u>) that enrolls Swiss residents of any age with confirmed or clinical diagnoses of PCD [22]. CH-PCD was established in 2013 and provides data for national and international PCD monitoring and research [23–27]. After a pilot period at the University hospital in Bern, it received approval nationwide from the Cantonal Ethics Committee of the Canton of Bern in 2015 (KEK-BE: 060/2015). Participants aged 14 years or older or parents of participants younger than age 14 provide written informed consent that allows collection of routinely available clinical data from hospitals or practices and invitations to participate in health-related questionnaire surveys.

For our analysis, we linked data on patient reported symptoms collected through a questionnaire survey to spirometry data from records available in the CH-PCD. We included all participants with a completed questionnaire and available lung function data from clinical follow-up within one year of completing the questionnaire. All French-or German-speaking participants registered in CH-PCD with a valid postal address received the questionnaire. We excluded one patient who spoke Italian and three patients with invalid postal addresses. We sent the questionnaire by post to all eligible participants in June 2022. Participants who had not yet responded received two postal reminders, one three weeks after the original questionnaire mailing and a second end of August 2022. Our reporting complies with the Strengthening the Reporting of Observational Studies in Epidemiology (STROBE) reporting recommendation [28].

### Questionnaire

The questionnaire was an adapted version of the FOLLOW-PCD patient questionnaire, part of the FOLLOW-PCD standardised tool for data collection for research and clinical follow-up [29]. The questionnaire included several questions about the frequency and characteristics of upper and lower respiratory, ear, and general symptoms, questions about lifestyle, and questions related to treatments, including having professional physiotherapy. There were three age-specific versions of the questionnaire: a) a parent/caregiver version for children aged 0-13 years, b) an adolescent version for those aged 14–17 years, and c) an adult version for participants aged ≥18 years. Most questions had predefined answer categories, and we included free text fields so participants could add any relevant comments. The FOLLOW-PCD questionnaire is available in German and French. Any additional questions were originally developed in German then translated into French by a native French speaker with knowledge about PCD.

### Patient-reported symptoms

The questionnaire asked about symptoms during the past three months. For our analysis, we focused on nasal symptoms, snoring, cough, sputum production, wheeze, shortness of breath, and chest pain. Questions about symptom frequency included five possible response categories: daily, often, sometimes, rarely, and never. We defined symptoms reported often or daily as frequent and symptoms reported sometimes or rarely as infrequent. In case of a missing reply, we classified the symptom as never reported.

### Demographic, diagnostic, and spirometry information

Sex, date of birth, area of residence, results of spirometry, and PCD diagnostic tests were retrieved from the CH-PCD database. We calculated age based on the questionnaire completion date. CH-PCD includes patients diagnosed following different algorithms, depending on the year when they were diagnosed because diagnostic tests for PCD and their availability have evolved over the years. All participants had a strong clinical suspicion for PCD and were managed accordingly by their respective physicians. For most patients, PCD diagnosis was based on a combination of tests in accordance with current recommendations, however, the diagnostic algorithm was not completed in all, and some were diagnosed based on strong clinical suspicion (*e.g.* Kartagener syndrome). Using the new unified diagnostic guidelines of the European Respiratory Society (ERS) and the American Thoracic Society (ATS), we defined confirmed PCD as two pathogenic or likely pathogenic variants in a known gene associated with PCD or a class I defect identified by transmission electron microscopy (TEM) [30, 31]. We defined as suspected PCD patients who had either not been tested with TEM or genetics or who had negative results for these tests and had one or more of the following: abnormal beat pattern in high frequency video microscopy; low (≤ 77 nL/min) nasal nitric oxide; pathologic immunofluorescence staining. Patients with a class 2 defect in TEM, genetic test result concordant with a known genotype-phenotype association, and one or more positive test results among nasal nitric oxide, high speed video microscopy, and immunofluorescence were classified as highly likely for PCD.

Lung function was measured by spirometry performed at the clinic or practice where participants were followed-up, by trained personnel according to ERS/ATS technical standards and local standard operating practices[32]. The assessment of the quality of spirometry was performed locally and we included patients younger than 6 years if the measurement was deemed well performed. We checked the data for plausibility, and calculated z-scores of FEV1, forced vital capacity (FVC), and of the FEV1/FVC ratio adjusted for age, sex, height and ethnicity, using the Global Lung Function Initiative 2012 reference values [33]. For each participant, one measurement was included in the analysis. If multiple measurements were available within the timeframe of one year from the questionnaire completion, we chose the measurement with the shortest time interval from the questionnaire. If pre- and post-bronchodilator measurements were available, we selected the one prior to the use of bronchodilators.

### Statistical analysis

We described population characteristics and reported frequency of respiratory symptoms, overall and by age group using median and interquartile range (IQR) for continuous variables and numbers and proportions for categorical variables. Missing data were reported as such. We assessed differences in basic characteristics between survey responders and non-responders, and between participants included and not included in the final analysis due to availability of lung function measurements using Pearson’s Chi square test. We assessed frequency of respiratory symptoms as potential determinant of lung function, namely FEV1 z-score, FVC z-score, and FEV1/FVC z-score, using multivariable linear regression models, separate for each symptom, adjusting each model for age, sex, and regular physiotherapy with a professional physiotherapist. To allow for graphical presentation, we ran our models again categorising age in the following categories: 0-13 years, 14-30 years, 31-50 years, and over 50 years. We performed all analyses using Stata version 16 (StataCorp LLC, Texas, USA).

## RESULTS

Sixty four out of 99 invited persons returned the questionnaire (response rate 65%), 54 of them had available spirometry data within one year period from the questionnaire completion (Figure 1). The median age was 24 years (IQR: 15–47) with 36 adults and 18 adolescents and children participating (Table 1). Most participants lived in German-speaking Swiss cantons (78%) and27 (50%) were female. When we compared characteristics of responders to non-responders as well as of people with available spirometry data to those without, we found no differences, except from a much higher proportion of females among the people who were excluded due to lack of spirometry data (Supplementary Tables S1 and S2).

**Figure 1:**
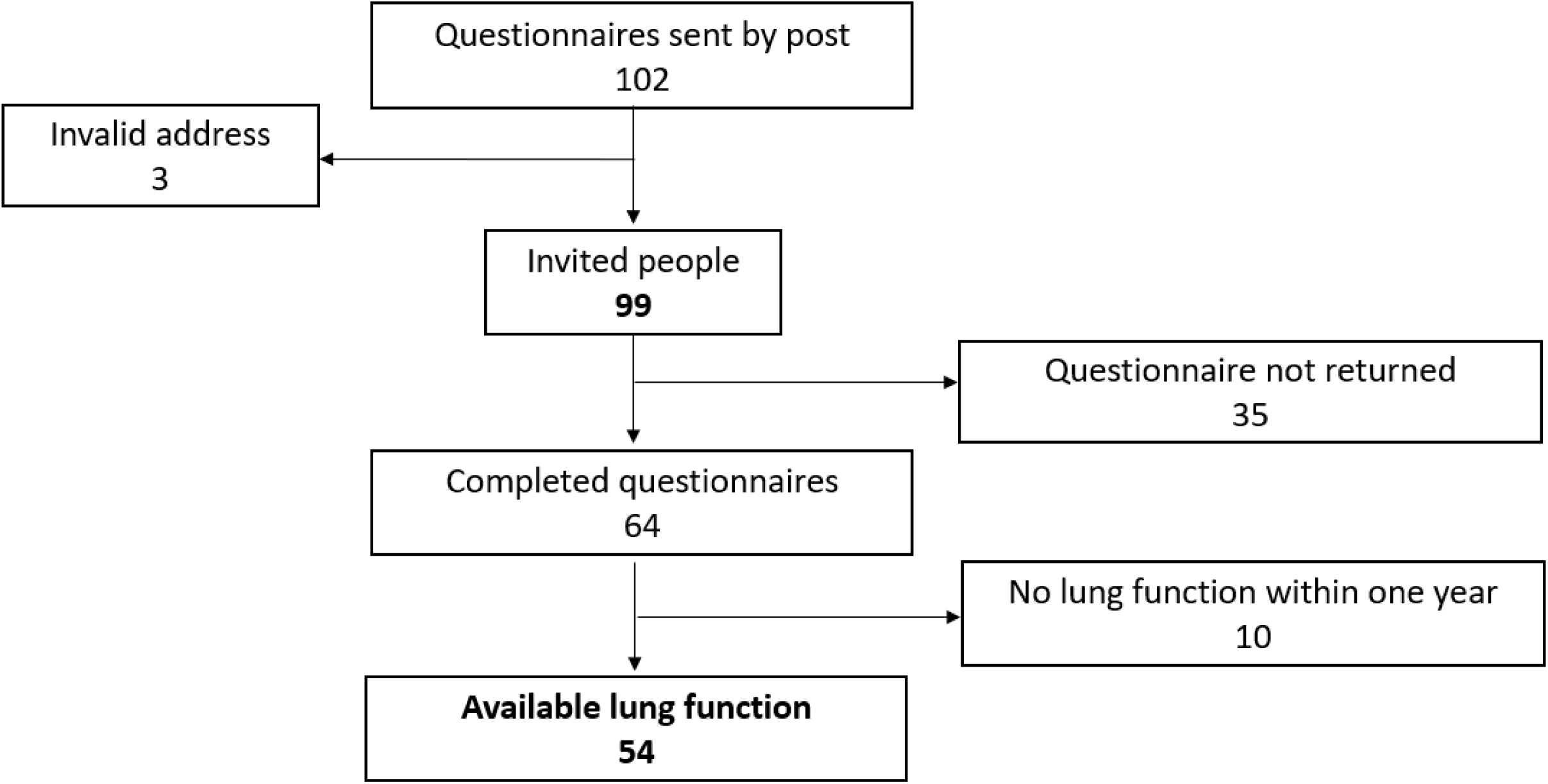
Flow chart of study participants with PCD living in Switzerland (Swiss PCD registry survey 2022).

**Table 1:** Characteristics of study participants with PCD living in Switzerland, overall and by age group (N=54).

|  | **N Total (%)** | **Children <18 y N (%)** | **Adults ≥18 y N (%)** |
| --- | --- | --- | --- |
| **Number of participants** | 54 (100) | 18 (100) | 36 (100) |
| **Age (years), median (IQR)** | 24 (15 – 47) | 13 (6 – 15) | 35 (24 – 55) |
| **Sex, female** | 27 (50) | 7 (39) | 20 (56) |
| **Area of residence** |  |  |  |
| German-speaking Switzerland | 42 (78) | 14 (78) | 28 (78) |
| French-speaking Switzerland | 12 (22) | 4 (22) | 8 (22) |
| **Diagnostic certainty#** |  |  |  |
| Confirmed PCD | 29 (54) | 12 (67) | 17 (47) |
| Highly likely PCD | 2 (4) | 1 (5) | 1 (3) |
| PCD suspected | 23 (42) | 5 (28) | 18 (50) |
| **Laterality defect** |  |  |  |
| Situs inversus | 13 (24) | 5 (28) | 8 (22) |
| Situs ambiguous | 1 (2) | 0 (0) | 1 (3) |
| Normal situs | 25 (46) | 10 (55) | 15 (42) |
| Not reported | 15 (28) | 3 (17) | 12 (33) |
| **Congenital heart defect** | 2 (4) | 1 (6) | 1 (3) |
| **Retinitis pigmentosa** | 1 (2) | 0 (0) | 1 (3) |
| **Hydrocephalus** | 1 (2) | 0 (0) | 1 (3) |
| **Tobacco exposure** |  |  |  |
| Active or ex-smoker | 7 (13) | 0 (0) | 7 (20) |
| Passive smoking (in household) | 5 (9) | 2 (11) | 3 (8) |
| No active or passive smoking | 42 (78) | 16 (89) | 26 (72) |
| **Occupation/school** |  |  |  |
| Student/preschool | 23 (43) | 18 (100) | 5 (14) |
| 81-100% employment | 17 (31) | 0 (0) | 17 (47) |
| 61-80% employment | 2 (4) | 0 (0) | 2 (6) |
| 41-60% employment | 3 (5) | 0 (0) | 3 (8) |
| ≤40% employment | 2 (4) | 0 (0) | 2 (6) |
| Retired/disability pension | 7 (13) | 0 (0) | 7 (19) |
| **Professional physiotherapy** |  |  |  |
| Yes | 21 (39) | 11 (61) | 10 (28) |
| No/not reported | 33 (61) | 7 (39) | 26 (72) |

PCD: Primary ciliary dyskinesia. y: years. All characteristics are presented as N and column % except for age in median and IQR: interquartile range. #: According to the ERS/ATS guidelines for the diagnosis of PCD- confirmed PCD: two pathogenic or likely pathogenic variants in a known gene associated with PCD or a class I defect identified by transmission electron microscopy (TEM), suspected PCD: patients who had either not been tested with TEM or genetics or who had negative results for these tests and had one or more of the following: abnormal beat pattern in high frequency video microscopy; low ( $\leq 77$ nL/min) nasal nitric oxide; pathologic immunofluorescence finding.

### PCD diagnosis and general characteristics

More than half of the participants (54%) had confirmed PCD according to the new ERS/ATS diagnostic guidelines [30]. Two patients had highly likely PCD based on class 2 defect identified with TEM, a test result concordant with known genotype-phenotype association, and one or more other positive diagnostic test results (Table 1). The remaining 23 (42%) participants were classified as suspected PCD. These patients had a strong clinical suspicion, usually typical Kartagener syndrome, and often one or more positive diagnostic test results, however, they had not completed fully the diagnostic algorithm, which is common in Switzerland as genetic testing for PCD was rare until recently and is still not performed routinely [34].

Twenty-five participants (46%) had normal situs and two a recorded congenital heart defect. Retinitis pigmentosa and hydrocephalus were only reported by one adult patient each. Seven adults reported occasional or frequent tobacco smoking or past smoking, and five additional participants (9%) reported passive smoking with household exposure. Seven adults were retired or on a disability pension. Twenty-one participants (39%) reported collaboration with a professional physiotherapist for their airway clearance.

### Respiratory symptoms

All participants reported chronic nasal symptoms, usually rhinorrhoea or obstruction, during the past three months, 46 (85%) of them frequently (Table 2). Snoring was less common with only three participants (6%) reporting it frequently and 26 (48%) infrequently. Among the lower respiratory symptoms, cough and sputum production were the most commonly reported items with 46 (85%) and 45 (83%) of participants reporting them frequently. Fifteen participants reported frequent wheeze (28%) and almost half (48%) of them infrequent. Shortness of breath was quite common, reported frequently by 16 (29.5%) of participants, 15 of whom were adults, while chest pain was only reported frequently by 6 (11%) of participants (Table 2). Missing responses to individual symptom questions were less than 5%.

**Table 2:**
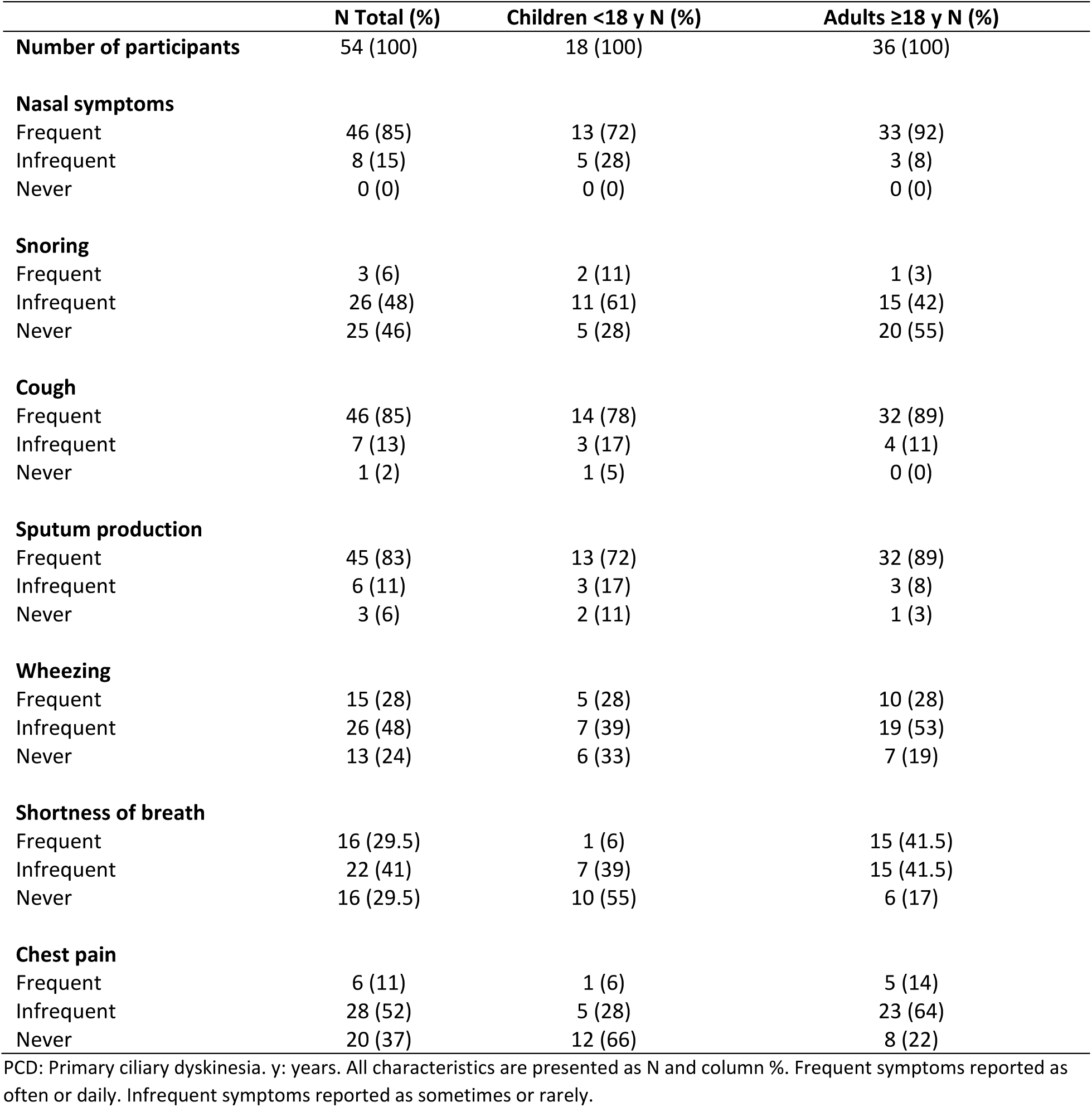
Respiratory symptoms of study participants with PCD living in Switzerland, overall and by age group (N=54).

|  | **N Total (%)** | **Children <18 y N (%)** | **Adults ≥18 y N (%)** |
| --- | --- | --- | --- |
| **Number of participants** | 54 (100) | 18 (100) | 36 (100) |
| **Nasal symptoms** |  |  |  |
| Frequent | 46 (85) | 13 (72) | 33 (92) |
| Infrequent | 8 (15) | 5 (28) | 3 (8) |
| Never | 0 (0) | 0 (0) | 0 (0) |
| **Snoring** |  |  |  |
| Frequent | 3 (6) | 2 (11) | 1 (3) |
| Infrequent | 26 (48) | 11 (61) | 15 (42) |
| Never | 25 (46) | 5 (28) | 20 (55) |
| **Cough** |  |  |  |
| Frequent | 46 (85) | 14 (78) | 32 (89) |
| Infrequent | 7 (13) | 3 (17) | 4 (11) |
| Never | 1 (2) | 1 (5) | 0 (0) |
| **Sputum production** |  |  |  |
| Frequent | 45 (83) | 13 (72) | 32 (89) |
| Infrequent | 6 (11) | 3 (17) | 3 (8) |
| Never | 3 (6) | 2 (11) | 1 (3) |
| **Wheezing** |  |  |  |
| Frequent | 15 (28) | 5 (28) | 10 (28) |
| Infrequent | 26 (48) | 7 (39) | 19 (53) |
| Never | 13 (24) | 6 (33) | 7 (19) |
| **Shortness of breath** |  |  |  |
| Frequent | 16 (29.5) | 1 (6) | 15 (41.5) |
| Infrequent | 22 (41) | 7 (39) | 15 (41.5) |
| Never | 16 (29.5) | 10 (55) | 6 (17) |
| **Chest pain** |  |  |  |
| Frequent | 6 (11) | 1 (6) | 5 (14) |
| Infrequent | 28 (52) | 5 (28) | 23 (64) |
| Never | 20 (37) | 12 (66) | 8 (22) |

PCD: Primary ciliary dyskinesia. y: years. All characteristics are presented as N and column %. Frequent symptoms reported as often or daily. Infrequent symptoms reported as sometimes or rarely.

### Lung function

The median difference between the date of questionnaire completion and the date of lung function measurement was 2.2 months (IQR: 0.5-4.9). Participants had a median FEV1 z-score of −2.29 (−3.37 – −1.03), FVC z-score of −1.23 (−2.23 – −0.24), and FEV1/FVC z-score of −1.63 (−2.70 – −0.54), with adults having lower z-scores compared to children (Table 3).

**Table 3:** Spirometric indices of study participants with PCD living in Switzerland, overall and by age group (N=54).

|  | **N Total (%)** | **Children <18 y N (%)** | **Adults ≥18 y N (%)** |
| --- | --- | --- | --- |
| **Number of participants** | 54 (100) | 18 (100) | 36 (100) |
| **Age (years), median (IQR)** | 24 (15 – 47) | 13 (6 – 15) | 36 (24 – 55) |
| **Sex, female** | 27 (50) | 7 (39) | 20 (56) |
| **FEV1 z-score** | -2.29 (-3.37 – -1.03) | -1.12 (-2.54 – 0.02) | -2.50 (-3.63 – -1.54) |
| **FVC z-score** | -1.23 (-2.23 – -0.24) | -1.03 (-1.56 – 1.12) | -1.47 (-2.78 – -0.48) |
| **FEV1/FVC z-score** | -1.63 (-2.70 – -0.54) | -1.13 (-2.13 – -0.10) | -1.87 (-2.89 – -0.76) |

PCD: Primary ciliary dyskinesia. y: years. Characteristics are presented as N and column % or as median and IQR: interquartile range. Age at lung function measurement.

### Association of respiratory symptoms with lung function

Patients reporting any wheeze had lower spirometric indices, for frequent wheeze FEV1 −1.11 (95%CI −2.20 – −0.20) and FEV1/FVC z-scores −1.00 (−1.83 – −0.18); for infrequent wheeze FEV1 - 1.13 (−2.13 – −0.12) and FEV1/FVC −1.24 (−2.00 – −0.48) compared to those who reported no wheeze (Table 4 and Figure 2). Similarly, participants who reported frequent shortness of breath had lower FEV1 (−1.27 [−2.50 – −0.04]) and FEV1/FVC z-scores (−0.99 [−1.96 – 0.01]). We found no signs of association between reported nasal symptoms, snoring, and chest pain with FEV1 and FEV1/FVC. The estimated coefficients for reported cough and sputum suggested a potential negative association with FEV1 and FEV1/FVC, with a magnitude of −0.75 (−3.90 – 2.37) for frequent cough and FEV1 z-score. Similarly estimated coefficients for reported sputum, wheeze, and shortness of breath suggested a negative association also with FVC z-score, however with smaller magnitude (Table S3). Increasing age was in general a predictor of lower lung function z-scores in all models (e.g. −0.02 [−0.05 – −0.01] annual FEV1 z-score decrease accounting for reported wheeze). Patients who were supported by a professional respiratory physiotherapist appeared to have higher z-scores (e.g. 0.90 [−0.02 – 1.81] accounting for reported shortness of breath).

**Figure 2:**
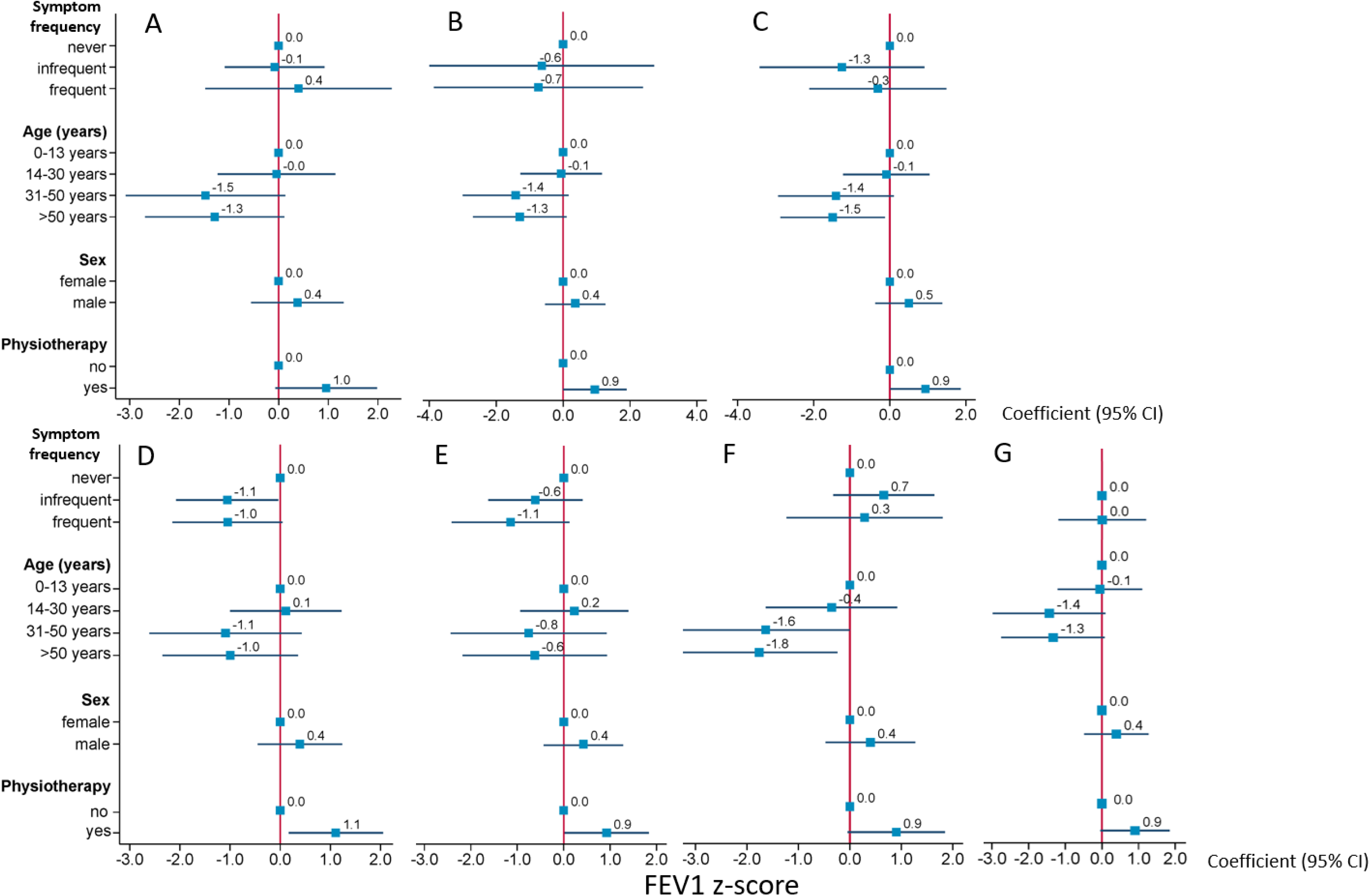
Determinants of FEV1 z-score among study participants with PCD living in Switzerland. A: snoring B: cough C: sputum D: wheezing E: shortness of breath F: chest pain G: nasal symptoms. z-scores are presented with 95% confidence intervals. Comparisons from baseline category, which is “never reported” for all symptoms, except for nasal symptoms where baseline is infrequent. Frequent symptoms reported as often or daily. Infrequent symptoms reported as sometimes or rarely.

**Table 4:** Multivariable linear regression analysis of spirometric indices in study participants with PCD living in Switzerland.

|  | FEV1 z-score |  |  | FEV1/FVC z-score |  |  |
| --- | --- | --- | --- | --- | --- | --- |
|  | Coefficient | 95% CIs | p-value<sup>#</sup> | Coefficient | 95% CIs | p-value<sup>#</sup> |
| **Nasal symptoms** (frequent) | 0.11 | -1.09 – 1.31 | 0.853 | 0.17 | -0.78 – 1.12 | 0.720 |
| Age | -0.03 | -0.05 – -0.01 | 0.022 | -0.02 | -0.04 – -0.01 | 0.021 |
| Male sex | 0.48 | -0.33 – 1.30 | 0.240 | 0.56 | -0.09 – 1.20 | 0.087 |
| Professional physiotherapy | 0.86 | -0.11 – 1.82 | 0.082 | 0.52 | -0.24 – 1.29 | 0.176 |
| **Snoring** (infrequent) | 0.11 | -0.86 – 1.07 | 0.828 | 0.16 | -0.60 – 0.93 | 0.669 |
| **Snoring** (frequent) | 0.38 | -1.48 – 2.25 | 0.680 | 0.03 | -1.44 – 1.51 | 0.965 |
| Age | -0.03 | -0.05 – -0.01 | 0.022 | -0.02 | -0.04 – -0.01 | 0.022 |
| Male sex | 0.50 | -0.32 – 1.36 | 0.219 | 0.60 | -0.06 – 1.27 | 0.076 |
| Professional physiotherapy | 0.84 | -0.23 – 1.90 | 0.065 | 0.47 | -0.37 – 1.31 | 0.263 |
| **Cough** (infrequent) | -0.70 | -3.99 – 2.60 | 0.673 | -0.77 | -3.31 – 1.77 | 0.544 |
| **Cough** (frequent) | -0.75 | -3.90 – 2.37 | 0.626 | -1.40 | -3.82 – 1.02 | 0.250 |
| Age | -0.03 | -0.05 – -0.01 | 0.028 | -0.02 | -0.04 – -0.01 | 0.037 |
| Male sex | 0.47 | -0.36 – 1.29 | 0.264 | 0.54 | -0.10 – 1.18 | 0.094 |
| Professional physiotherapy | 0.92 | -0.05 – 1.89 | 0.062 | 0.67 | -0.07 – 1.42 | 0.076 |
| **Sputum** (infrequent) | -1.44 | -3.57 – 0.70 | 0.182 | -0.65 | -2.37 – 1.08 | 0.454 |
| **Sputum** (frequent) | -0.41 | -2.20 – 1.38 | 0.647 | -0.36 | -1.81 – 1.08 | 0.616 |
| Age | -0.03 | -0.05 – -0.01 | 0.010 | -0.02 | -0.04 – -0.01 | 0.022 |
| Male sex | 0.58 | -0.23 – 1.38 | 0.155 | 0.61 | -0.05 – 1.26 | 0.068 |
| Professional physiotherapy | 0.90 | -0.05 – 1.85 | 0.061 | 0.59 | -0.18 – 1.35 | 0.129 |
| **Wheeze** (infrequent) | -1.13 | -2.13 – -0.12 | 0.029 | -1.24 | -2.00 – -0.48 | 0.002 |
| **Wheeze** (frequent) | -1.11 | -2.20 – -0.20 | 0.046 | -1.00 | -1.83 – -0.18 | 0.018 |
| Age | -0.02 | -0.05 – -0.01 | 0.048 | -0.02 | -0.33 – -0.01 | 0.062 |
| Male sex | 0.45 | -0.33 – 1.22 | 0.250 | 0.53 | -0.05 – 1.12 | 0.074 |
| Professional physiotherapy | 1.07 | 0.11 – 2.02 | 0.028 | 0.81 | 0.10 – 1.53 | 0.027 |
| **Shortness of breath** (infrequent) | -0.67 | -1.65 – 0.31 | 0.174 | -0.53 | -1.31 – 0.24 | 0.174 |
| **Shortness of breath** (frequent) | -1.27 | -2.50 – -0.04 | 0.043 | -0.99 | -1.96 – 0.01 | 0.048 |
| Age | -0.01 | -0.04 – 0.01 | 0.294 | -0.01 | -0.03 – -0.01 | 0.297 |
| Male sex | 0.46 | -0.33 – 1.24 | 0.250 | 0.54 | -0.08 – 1.17 | 0.086 |
| Professional physiotherapy | 0.90 | -0.02 – 1.81 | 0.054 | 0.57 | -0.15 – 1.29 | 0.120 |
| **Chest pain** (infrequent) | 0.70 | -0.21 – 1.62 | 0.129 | -0.03 | -0.77 – 0.72 | 0.944 |
| **Chest pain** (frequent) | 0.25 | -1.17 – 1.66 | 0.727 | -0.04 | -1.19 – 1.11 | 0.944 |
| Age | -0.03 | -0.57 – - 0.01 | 0.008 | -0.21 | -0.41 – 0.00 | 0.033 |
| Male sex | 0.50 | -0.30 – 1.31 | 0.216 | 0.57 | -0.86 – 1.23 | 0.087 |
| Professional physiotherapy | 0.90 | -0.05 – 1.85 | 0.064 | 0.56 | -0.21 – 1.33 | 0.152 |

Results of multivariable regression models separate for each symptom. PCD: primary ciliary dyskinesia. CI: confidence interval. FEV1: Forced Expiratory Volume in 1 second. FVC: Forced Vital Capacity. Age measured in years #: comparisons from baseline category, which is “never reported” for all symptoms, except for nasal symptoms where baseline is infrequent. Frequent symptoms reported as often or daily. Infrequent symptoms reported as sometimes or rarely.

## DISCUSSION

To our knowledge this is the first study that assesses associations of spirometry, the most commonly used outcome measure for chronic respiratory diseases, with the broad range of respiratory symptoms patients with PCD experience. Patients with PCD living in Switzerland reported a variety of symptoms, many of them experienced frequently, and had low lung function z-scores even in childhood. Among all reported respiratory symptoms, wheeze and shortness of breath were associated with lower FEV1 and FEV1/FVC. Despite the lack of statistical significance, the coefficient’s size for other lower respiratory tract symptoms such as cough and sputum, suggested still a negative association with FEV1 and FEV1/FVC. Upper airway symptoms showed no signs of association with spirometry.

The most important strength of our study was that it was nested in the CH-PCD allowing to invite all registered participants currently in follow-up living in Switzerland without any selection. It also allowed to link patient-reported information on symptoms to clinical data on lung function retrieved from clinics and practices. Another strength was the use of the standardised FOLLOW-PCD patient questionnaire [29], which led to homogeneous, standardised data about symptom frequency reported directly from participants or parents of participants.

Our study presents several limitations. Not all invited participants completed the questionnaire and among those who did, not all had lung function measurements available. Although we did not find many differences in baseline characteristics when comparing these groups, this could have potentially introduced selection bias as people with more severe symptoms might be more prone to participate. Similarly, people with less severe disease could be more prone to have missed their scheduled appointment for lung function measurement. It would be relevant to distinguish between participants with an additional asthma diagnosis, but this information was not regularly recorded in clinical records. Moreover, reversible airway obstruction appears to be an independent feature of PCD, which is often misdiagnosed as asthma [35] so information on asthma diagnosis is not always accurate. Although national, CH-PCD still has few registered participants as PCD remains underdiagnosed. Our sample size is reasonable for a rare disease in a small country, it led, however, to limitations in considering other important factors in our analysis, such as exposure to tobacco, environmental, or further clinical factors. Our findings on reported symptoms are similar to previous studies. In 2020, we had sent another survey to CH-PCD participants with the same questions on symptoms as it was also based on the FOLLOW-PCD patient questionnaire. Symptom frequency was similar overall with higher prevalence of frequent wheeze (28% in 2022 versus 12% in 2020) and frequent shortness of breath (29.5% versus 20%), which could be due to the fact that participants seemed to experience less symptoms during the COVID pandemic, probably due to shielding and less infections [23]. Compared to an international study assessing the prevalence of sinonasal problems among patients with PCD, our participants reported more frequent nasal symptoms (52% versus 85%) but less frequent snoring (12% versus 6%) perhaps caused by different age distribution [27]. Other studies reporting on clinical symptoms use mostly nonstandard information from clinical charts and show a high variability in reported symptoms, not easy to compare with our results.

There is only one study that compares how well reported symptoms correlate to objective measures among patients with PCD, focusing on upper airway and ear manifestations [36]. In this study analysing international data, we found that many patients appeared to underestimate and underreport their symptoms, to which they grew accustomed over time, and that patient-reported measures complement objective measures. The current study’s findings also highlight that objective measures cannot replace information on patient-reported symptoms, supporting the use of both for research and clinical follow-up.

Although universally accepted and commonly used, spirometry has shown to be not sufficient on its own for monitoring respiratory morbidity in PCD. Longitudinal spirometry-derived lung function measurements from an international prospective cohort showed that changes in intra-individual FEV1 between study visits in stable disease conditions were beyond the expected test variability [37]. Compared to spirometry, multiple breath washout has been shown to be more sensitive in identifying early lung damage, and could be used additionally to guide appropriate management especially in patients diagnosed early in life [16, 38–40]. Moreover, discordance between lung function and imaging outcomes in patients with PCD supports the use of both imaging and lung function [17]. Regular standardised assessments such as nasal endoscopy and sinus imaging are required to evaluate how upper airway disease progresses [27].

Spirometry captures some of the lower airway symptoms, but cannot capture the range of respiratory symptoms patients with PCD experience in daily life. For research and clinical follow-up, we need additional objective outcome measures e.g. assessing lung clearance index, imaging outcomes, and upper airway assessments, together with regular and standardised assessment of patient-reported outcomes to successfully assess disease progression and guide management.

## Author Contributions

M Goutaki developed the concept and designed the study. M Goutaki, M Koller, YT Lam, and L Schreck organised the survey, then cleaned and standardised the data. M Goutaki, YT Lam, and M Koller performed the statistical analyses and drafted the manuscript.

All authors commented and revised the manuscript. M Goutaki takes final responsibility for content.

## Data sharing statement

CH-PCD study data is not deposited at an open access repository. Because of the rarity of the disease although data is pseudonymised, it may include still sensitive information; therefore, participants were not asked to give consent to have their data deposited publicly. Partial datasets for specific analyses including individual patient data and a data dictionary defining each included field can be available from Dr Goutaki upon reasonable request.

## Acknowledgments

We want to thank all the people with PCD in Switzerland and their families for participating in this survey and in the CH-PCD. We are also grateful to the Swiss PCD support group that closely collaborates with us. We would like to acknowledge all members of the current extended Swiss PCD research group for their support to the CH-PCD. The current Swiss PCD research group includes (in alphabetical order): Juerg Barben (Children’s Hospital of Eastern Switzerland St. Gallen), Sylvain Blanchon (University Hospital of Lausanne), Jean-Louis Blouin (University Hospital of Geneva), Marina Bullo (University Hospital of Bern), Carmen Casaulta (University Hospital of Bern), Christian Clarenbach (University Hospital of Zurich), Regula Corbelli (University Hospital of Geneva), Andrea Fernandez Rodriguez (ISPM, University of Bern), Vasiliki Gkatzou (ISPM, University of Bern), Myrofora Goutaki (ISPM, University of Bern), Nicolas Gürtler (University Hospital of Basel), Beat Haenni (Institute of Anatomy, University of Bern), Andreas Hector (Cantonal Hospital Winterthur), Michael Hitzler (Children’s Hospital Lucerne), Andreas Jung (Cantonal Hospital Winterthur & University Children’s Hospital Zurich), Lilian Junker (Hospital Thun), Peter Iseli (Cantonal Hospital Chur), Nena Karavasiloglou (ISPM, University of Bern), Elisabeth Kieninger (University Hospital of Bern), Claudia E. Kuehni (ISPM, University of Bern), Yin Ting Lam (University Children’s Hospital Zurich), Philipp Latzin (University Hospital of Bern), Romain Lazor (University Hospital of Lausanne), Dagmar Lin (University Hospital of Bern), Marco Lurà (Children’s Hospital Lucerne), Loretta Müller (University Hospital of Bern), Nicolas Regamey (Children’s Hospital Lucerne), Isabelle Rochat (University Hospital of Lausanne), Daniel Schilter (Quartier Bleu Bern), Iris Schmid (Quartier Bleu Bern), Valérie Schwartz (ISPM, University of Bern), Bernhard Schwizer (Quartier Bleu Bern), Elias Seidl (University Children’s Hospital Zurich), Andrea Stokes (University Hospital of Bern), Daniel Trachsel (University Children’s Hospital Basel), Stefan A. Tschanz (Institute of Anatomy, University of Bern), Johannes Wildhaber (University of Fribourg), and Maura Zanolari (Hospital of Bellinzona).

---

**Table S1:** Comparison of demographic characteristics of CH-PCD participants who responded to the survey to non-responders.

|  | **Invited N (%)** | **Responded to survey**<br>**N (%)** | **Non-responders**<br>**N (%)** | **p-value<sup>§</sup>** |
| --- | --- | --- | --- | --- |
| **Number of participants** | 102 (100) <sup>#</sup> | 64 (100) | 38 (100) <sup>#</sup> |  |
| **Age, median (IQR)** | 24 (2-75) | 24 (3-75) | 25 (2-71) |  |
| **Female sex** | 54 (53) | 35 (55) | 19 (50) | 0.647 |
| **German speaking** | 80 (78) | 49 (77) | 31 (82) | 0.551 |

PCD: primary ciliary dyskinesia. CH-PCD: Swiss PCD registry. All characteristics are presented as N and column % except for age in median and IQR: interquartile range. #: includes 3 participants with an invalid address §: Pearson's Chi square test

**Table S2:** Comparison of demographic characteristics of participants included in the analysis compared to the ones who were excluded because of no available lung function measurement within the defined period.

|  | **Survey responders**<br>**N (%)** | **Participants included in analysis**<br>**N (%)** | **Participants excluded (no spirometry)**<br>**N (%)** | **p-value<sup>§</sup>** |
| --- | --- | --- | --- | --- |
| **Number of participants** | 64 (100) | 54 (100) | 10 (100) |  |
| **Age, median (IQR)** | 22 (3-73) | 22 (3-73) | 28 (3-65) |  |
| **Female sex** | 35 (55) | 27 (50) | 8 (80) | 0.080 |
| **German speaking** | 49 (77) | 40 (74) | 9 (90) | 0.275 |

PCD: primary ciliary dyskinesia. CH-PCD: Swiss PCD registry.
All characteristics are presented as N and column % except for age in median and IQR: interquartile range. §: Pearson's Chi square test

**Table S3:**
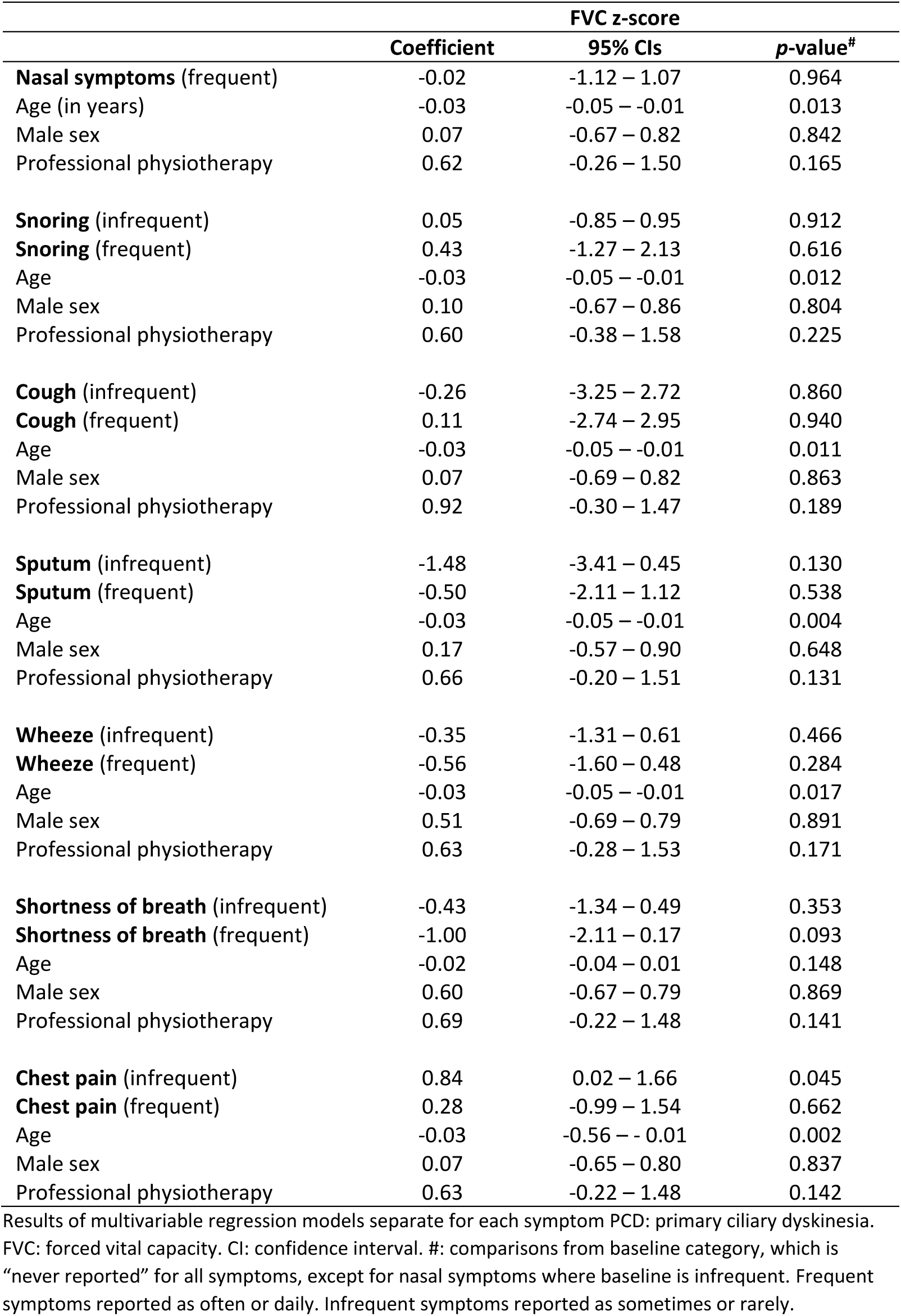
Multivariable linear regression analysis of FVC z-score in study participants with PCD living in Switzerland.

